# Perineal and rectal nerve recruitment order varies during pudendal neurostimulator implant surgery

**DOI:** 10.1101/2024.07.13.24310332

**Authors:** Po-Ju Chen, Amador C. Lagunas, Vanessa Soriano, Priyanka Gupta, Tim M. Bruns

## Abstract

**Introduction:** Pudendal nerve stimulation (PNS) is an off-label therapy for patients experiencing pelvic pain and voiding dysfunction. The pudendal nerve has two efferent branches to the rectum and perineum. Only the rectal branch is monitored via external anal sphincter electromyography during the implant procedure to help determine the lead position. We examined intraoperative PNS-driven urethral pressures to infer nerve recruitment order and tracked patient reported outcomes.

**Methods:** Patients receiving PNS for pelvic pain and/or urinary symptoms were recruited. During the implant surgery, urethral pressure was measured with a multi-sensor pressure catheter placed in the lower urinary tract. PNS-driven changes in urethral pressure and external anal sphincter (EAS) electromyography were compared to determine the relative perineal and rectal nerve recruitment order. Participants completed pelvic pain, bladder, bowel, and sexual function surveys before and after the surgery. The primary outcome measure was the relative nerve recruitment order during PNS. Secondary outcome measures were the PNS-driven urethral responses and changes in survey symptom scores due to PNS.

**Results:** Data was collected from thirteen intraoperative sessions. Seven participants had rectal nerve recruitment first, four participants had perineal nerve recruitment first, and two participants had mixed nerve recruitment during intraoperative PNS. The average normalized urethral pressure change was 4.7% at the EAS threshold, 59.2% at twice EAS threshold, and 68.2% at three times the EAS threshold. Urethral pressure changes for each participant often varied between different active PNS electrodes. Participants had significant improvements in pelvic pain and bladder function survey scores with PNS (p < 0.04). There was no relationship between nerve recruitment order and changes in any surveys.

**Conclusion:** PNS can recruit the perineal nerve before the rectal nerve. Each lead electrode may trigger different urethral response patterns within a participant. This study provided new insights into the effect of PNS on the recruitment of nerves in the pelvis and may help guide future surgical placement of PNS systems.

## Introduction

Pudendal nerve stimulation (PNS) is an off-label use of sacral neuromodulation (SNM) for targeting pelvic pain and bladder symptoms^1-3^. Studies have shown that patients whose symptoms are refractory to SNM can be salvaged with PNS leading to improvement of pelvic pain, voiding, and urinary incontinence^4,5^. However, there is limited literature on the direct effects of PNS on the lower urinary tract^6,7^, in particular during intraoperative placement.

The pudendal nerve originates from the S2, S3, and S4 spinal nerve roots. It typically forms a primary trunk and then splits into the inferior rectal nerve, perineal nerve, and genital nerve primary branches^8-10^. The genital nerve, which is sensory only, innervates the external genitalia. The perineal nerve innervates pelvic floor, urethra, and perineal muscles, while the rectal nerve innervates the external anal sphincter (EAS) and levator ani^11^. Pudendal nerve anatomical structure is diverse^8,12^. Near the sacrospinous ligament, one to three nerve trunks may be found with different branching points^12^. The heterogeneity of pudendal nerve anatomy could pose potential obstacles during a PNS surgical approach and may influence the lead placement and variation in results. No study has investigated the activation patterns of the pudendal nerve among perineal and rectal motor responses during PNS implant surgery.

The PNS implant typically involves two stages of outpatient surgery, following standard SNM procedures. During surgical access to the pudendal nerve in the stage-1 implant procedure, patients are prone and sedated while surgeons use anatomical landmarks, fluoroscopic guidance, and EAS electromyography (EMG) to place the quadripolar lead^3,7,13^. The EAS EMG monitoring only accounts for the rectal nerve motor responses, neglecting the perineal nerve. An optimal lead position is parallel to the main pudendal trunk about 5 mm away, with at least 2 electrodes consistently eliciting EAS contractions at low stimulation amplitude without causing leg movements. After approximately 2 weeks if the patient’s primary symptoms are reduced by at least 50% an implantable pulse generator will be implanted during a stage-2 surgery.

In this study, we employed a multi-sensor pressure catheter within the urethra during the stage-1 pudendal neurostimulator implant. This allowed us to examine changes in urethral pressure caused by stimulation and determine the relative order of pudendal nerve recruitment between the perineal and rectal nerves amongst participants. In this study we collected 13 intraoperative datasets from 12 female participants and compared pudendal nerve recruitment with clinical outcomes across this cohort.

## Materials & Methods

### Participant recruitment

Patients referred for PNS treatment were assessed for eligibility and contacted. Participants eligible for this study had to be at least 18 years old and capable of filling out questionnaires and interacting with the research team. Patients with certain medical conditions, including pregnancy, areflexive/atonic/ neurogenic bladder, or other conditions affecting the micturition neural circuits were excluded.

### Intra-operative set up

The PNS implant system received by participants in this study includes a 4-electrode lead (model 978B141 or 978A128 InterStim SureScan, Medtronic) and an implantable pulse generator (model 3058 recharge-free InterStim II, model 97810 rechargeable InterStim Micro, or model 97800 recharge-free InterStim X, Medtronic). The specifics of the pudendal neurostimulator implant procedure have been described previously^3,13^. The data collection for this study took place during the lead implant stage-1 surgery.

Each study participant underwent standard preparation for their stage-1 implant procedure with the addition of a multi-sensor manometry pressure catheter (Manoscan MSC-3886, Medtronic)^14^. The 2.75-mm diameter (8.25 French gauge) catheter had 36 circular sensors, each 4 mm in length with 7.5 mm spacing (center to center of two adjacent sensors). The catheter was inserted into the urethra to the bladder (Figure S1) when the participant was first positioned on the surgical table and was taped to the inside of one thigh to secure its location in the urethra. The catheter was connected to a high-resolution esophageal manometry system (Manoscan ESO, Medtronic) with a sampling rate of 100 data points per second per sensor.

After catheter insertion, the participant was placed and secured in a prone position on the surgical table. The experimental setup also included standard intraoperative EAS EMG monitoring procedures^13^. EMG needle electrodes (Ambu Neuroline, twisted pair subdermal, 74612-250/1/20) were inserted on either lateral side of the EAS for monitoring stimulation-triggered EMG responses.

The goal of the stage-1 surgery is to place the 4-electrode lead parallel to the main trunk of the pudendal nerve near Alcock’s canal, with EAS EMG responses visible in response to stimulation on at least two electrodes. The EAS motor threshold (EASt) was determined by the minimum amplitude required to trigger EAS EMG signals for each electrode. Following the placement of the neurostimulator lead, the EASt for each electrode was determined, and the final position of the lead was documented with fluoroscopy images (Figure S1).

### Intraoperative pudendal nerve stimulation

Stimulation trials were performed after the lead placement and before the final closure of incisions. Experimental stimulation was applied at 3 Hz to allow for the return of urethral pressure to pre-stimulation baselines before the next stimulus pulse. The InterStim system has a fixed pulse width range of 60 to 450 μs, with a standard value of 210 μs^15^. In this study, we primarily used a pulse width of 210 μs and performed limited trials at the device minimum (60 μs) and maximum (450 μs) pulse widths due to time constraints. The stimulation amplitude was adjusted based on multiple increments of the EASt for an electrode, up to a magnitude that did not induce discomfort or leg motion for patients, with a maximum limit of 5.5 mA. Each stimulation trial lasted for a minimum of 10 seconds, providing a period of stable urethral responses without artifacts. Only the data gathered during a stable period was quantified.

### Urethral pressure data analysis

The pressure catheter was operated on its manufacturing software (ManoScan acquisition software, Medtronic). The retrieved data was converted from a text format (.txt) into an Excel file format (Microsoft Excel, .xlsx) for analysis.

The relative position of the catheter with respect to the lower urinary tract was estimated by reviewing the location of the pubic symphysis and the EAS EMG needles around the anus in the intraoperative fluoroscopy image (Figure S1). The Manoscan pressure channel(s) with the highest or two similarly highest pressure readings during a period of no stimulation were identified as being in the region of the bladder neck/proximal urethra. The first sensor that was not at room pressure was identified as the distal urethra location.

For each trial, the 3 Hz frequency stimulation pattern was identified on each channel, if present. The mean and standard deviation for the pressure change (ΔP) in each stimulation trial was determined for the peak-to-peak pressure change across ten consecutive stimulation responses during a stable period when the stimulation amplitude was fixed and there were no signal artifacts. Trials without a 3 Hz peak-to-peak stimulation response were identified as having no response. For all participants except two (1008 and 1010), ΔP could be determined during periods with stable respiration artifacts.

Across participants, the peak-to-peak ΔP for each sensor in a given trial was normalized to the maximum pressure change (max ΔP) observed at that sensor. This was done to examine the relative stimulation-driven effect on the urethra in comparison to the increase in stimulation amplitude within a series that had the same stimulation parameters.

### Clinical outcomes analysis

The primary clinical outcome was determined by the participants’ decision to either retain the neurostimulator implant or have it removed within the timeframe of our study. Matching sets of self-assessment surveys related to pelvic organ function were collected before the stage-1 surgery and again at least two weeks after the stage-2 procedure. These surveys assessed pelvic pain symptoms (Female Genitourinary Pain Index – fGUPI)^16^, overall bladder symptoms (American Urological Association Symptom Index – AUASI)^17^, urinary incontinence (Michigan Incontinence Symptom Index – M-ISI)^18^, bowel symptoms (Colorectal-Anal Distress Inventory 8 – CRAD-8)^19^, and sexual function (Female Sexual Function Index – FSFI)^20^.

Survey outcomes were reported as the average and standard deviation of the total score across participants. We used a paired Student’s t-test to determine whether any surveys indicated significant changes in symptoms during the study. To evaluate whether the nerve recruitment order was correlated with symptom changes after PNS implantation, participants were grouped based on the first recruited nerve by the 4-electrode lead: the rectal nerve, the perineal nerve, or a combination of both rectal and perineal nerves (mixed). We calculated the 95% confidence interval for the change in each survey score based on the population standard deviation of the three nerve activation clusters due to the small group sizes.

## Results

We collected data from 12 participants who underwent 13 successful pudendal neurostimulator implant surgeries (Table 1). The majority of participants were suffering from pelvic pain with concurrent bladder issues. We omitted three male study participants to focus on female-participant responses in this report. The timeline for subject participation in this study is given in Figure S2.

**Table 1.**
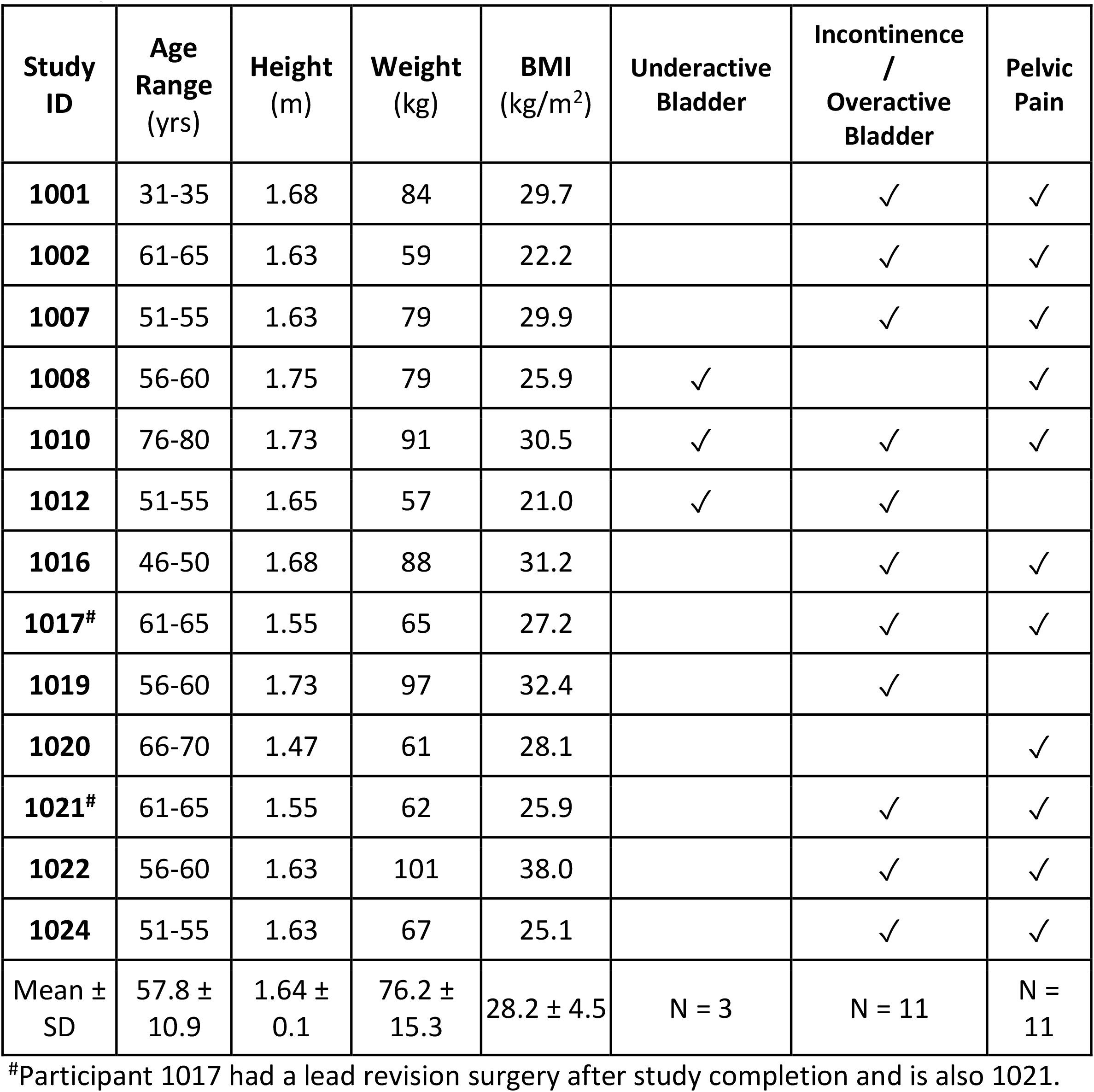
Study participant information. The participants were all Caucasian without Hispanic ethnicity.

The average participant baseline pressures during a 10-second stable period before stimulation trials were 25.4 ± 9.9 cmH_2_O in the bladder, 57.0 ± 13.7 cmH_2_O in the proximal urethra/bladder neck, and 19.7 ± 7.9 cmH_2_O in the distal urethra (Figure S3). There was a significant difference in the baseline pressure among lower urinary tract locations (one-way ANOVA; F-ratio 41.8125, p-value < 0.0001). The mean proximal urethra pressure was significantly different (t-test; p-value < 0.0001) than in the bladder and distal urethra. The highest pressure occurred in the bladder neck and proximal urethra in all participants, except one outlier (1008). The average maximum urethral closure pressure (MUCP, the difference between the maximum urethral pressure and bladder pressure) across participants was 31.6 ± 9.3 cmH_2_O.

Across participants, E0 and E1 generally had a lower EASt (1.1 ± 0.4 mA, 1.4 ± 1.0 mA respectively) than E2 and E3 (1.9 ± 1.1 mA, 2.1 ± 0.9 mA respectively). There were no differences in EASt between the electrodes (One-way ANOVA; F-ratio 2.56, p-value 0.068). E1 and E2 electrodes evoked EAS responses in all participants. In Table S1, EASt is given for each participant electrode, for monopolar 3 Hz, 210 µs stimulation.

Most of the participants had 4 pressure sensors in their functional urethra, except two participants (1001, 1024) who had 3 sensors. Eleven participants had measurable stimulation-driven urethral pressure changes. The maximum peak-to-peak pressure change (max ΔP) for each urethral sensor is given in Figure 1. Two participants (1008, 1010) did not have urethra responses within our testing range. All 11 responding participants had stimulation-driven responses in the proximal urethra (top two proximal urethra sensors). Three of these participants (1007, 1012, 1021) did not have responses at the distal site(s). The largest max ΔP per participant was driven by E0 (30%), E1 (43%), or E2 (17%). In two participants (1002, 1022) we observed an elevated pressure tone in the proximal urethra when stimulation was ramped up (Figure S4). There was no difference in max ΔP among urethral pressure sensor sites (One-way ANOVA; F-ratio 0.65, p-value 0.59).

**Figure 1.**
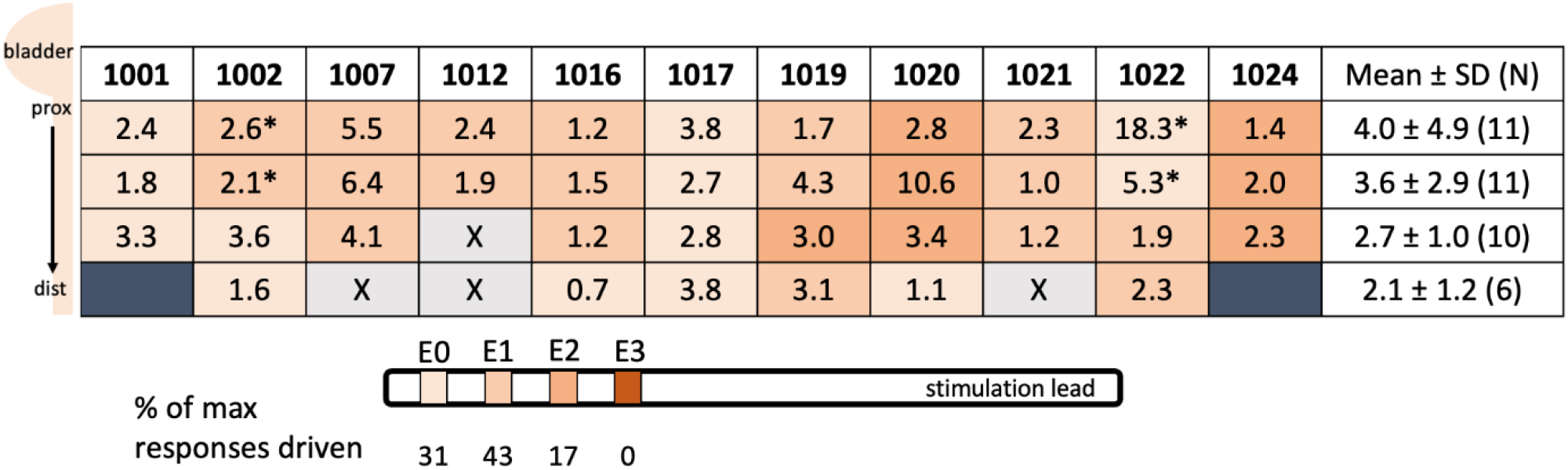
Maximum urethral peak-to-peak ΔP across the urethra in each participant. Darkened cells indicate participants with only 3 functional sensors in their urethra. Light grey cells with X represent functional sensors without urethral pressure responses. Asterisks indicate pressure tone elevation in addition to 3 Hz peak-to-peak pressure responses (see Figure S4). Color-coding indicates the electrode which was used to generate the pressure.

We investigated the effect of increases in stimulation amplitude with respect to EASt (Figure 2). Across participants at 1x EASt, we observed an average peak-to-peak ΔP normalized to the max ΔP of 4.7% across the urethral locations (2.3-7.1% per-position range). At 2x EASt the urethral responses consistently increased, with 11.9% reaching max ΔP. We observed 59.2% of the average normalized ΔP across the urethral locations at 2x EASt (52.2-67.9% per-position range). For 3x EASt and 4x EASt, most urethral responses increased (68.2-79.0% across urethra), however six participants had decreasing ΔP trends.

**Figure 2.**
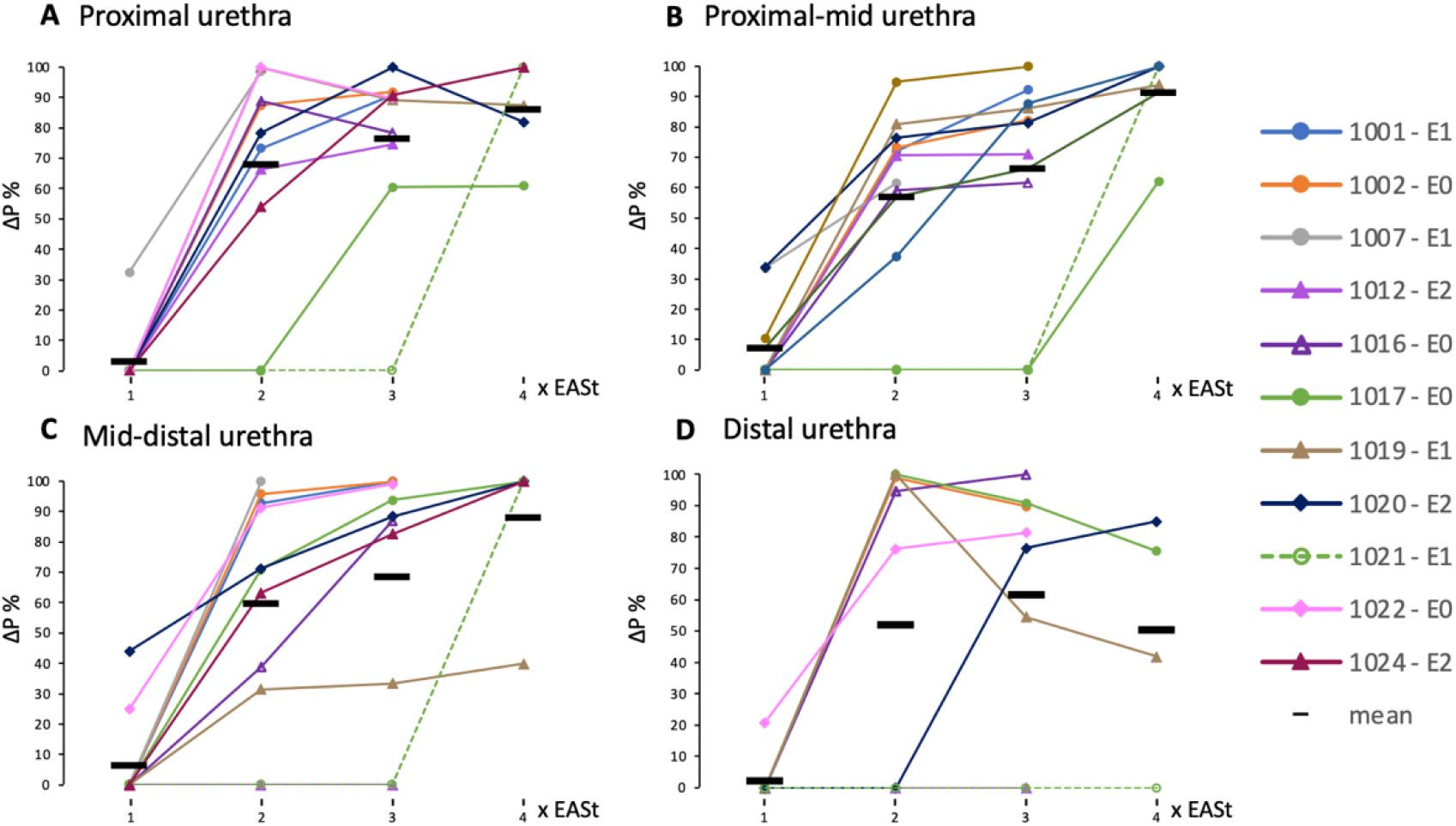
Normalized increases in urethral pressure per location for the most consistent electrode for each participant. The electrode that yielded the most per-location maximal responses (or overall maximum urethra response if multiple electrodes had equal number of per-location maximums) across the urethra was identified as the most consistent in each participant. Stimulation-driven peak-to-peak ΔP was normalized to the maximum urethral ΔP response per urethral sensor across all electrodes for a given participant. Horizontal bars indicate group means of the consistent electrode trials. The most consistent electrode for each participant is indicated in the legend.

The onset order of stimulation-driven urethral responses at different locations suggested the relative recruitment order of the distal pudendal branches. At a stimulation amplitude of 1x EASt, if urethral responses were already prominent, we classified it as having perineal nerve recruitment first (e.g., indicated by values above 0% ΔP at 1x EASt in Figure 2). Conversely, if no urethral responses were identified at 1x EASt then we classified it as having rectal nerve recruitment first. Two examples of nerve recruitment orders are given in Figure 3. The stimulation recruitment order was unique to individual participants, and different electrodes triggered various responses within a participant. Detailed information about the nerve recruitment order for each electrode among all participants is in Table S2.

**Figure 3.**
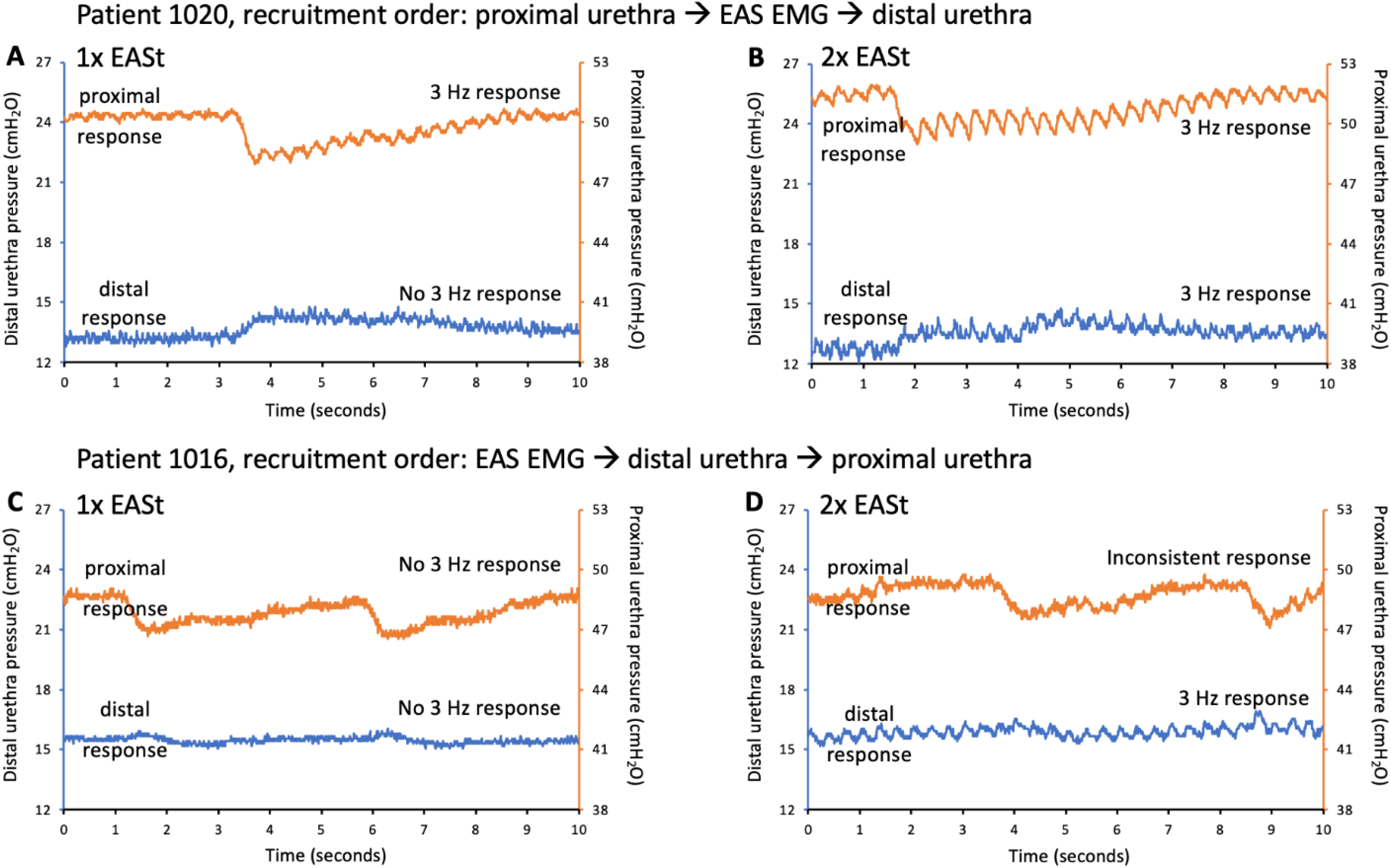
Nerve recruitment order examples. Participant 1020 had clear 3 Hz stimulation-driven responses at 1x EASt in the proximal urethra (A), and the distal urethra responses appeared at 2x EASt (B). For this participant, the proximal perineal nerve was recruited before the rectal nerve, with the distal perineal nerve recruited last. In participant 1016, there were no urethral responses at 1x EASt (C), and the distal urethra had clearer 3 Hz responses than the proximal urethra at 2x EASt (D). In this participant, the rectal nerve was the first recruited, followed by distal perineal and proximal perineal nerves. Each figure part represents the stable period for the given stimulation amplitude, with stimulation on for the full 10 seconds. The slow oscillation in the pressure tone, generally lasting 5-7 seconds per cycle, is likely due to changes in pelvic floor pressure during different phases of respiration.

The nerve recruitment orders that most accurately represent the participant activation patterns are summarized in Table 2. Seven participants had the rectal nerve recruited first. Two participants (1001, 1012) demonstrated mixed initial nerve recruitment (perineal or rectal nerve) in response to stimulation on different electrodes. Four participants (1007, 1017, 1020, and 1022) had either the proximal or distal perineal nerve recruited before the rectal nerve.

**Table 2.** Summary of nerve recruitment order, implant outcome, and pelvic function surveys. Zero survey scores indicate no dysfunction except FSFI, for which a score of 36 indicates no dysfunction.

| ID | Nerve Recruitment Order | Outcome | Pain<br>fGUIPI<br>(0 to 45) |  | Incontinence<br>M-ISI<br>(0 to 40) |  | Bladder<br>AUASI<br>(0 to 35) |  | Bowel<br>CRAD8<br>(0 to 100) |  | Sexual Function<br>FSFI<br>(36 to 0) |  |
| --- | --- | --- | --- | --- | --- | --- | --- | --- | --- | --- | --- | --- |
|  |  |  | Pre | Post | Pre | Post | Pre | Post | Pre | Post | Pre | Post |
| 1001 | Distal PER / REC < Proximal PER | explant | 27 | 17 | 1 | 2 | 8 | 2 | 34.4 | 43.8 | 5.0 | 7.5 |
| 1002 | REC < Distal PER = Proximal PER | positive | 21 | 5 | 16 | 8 | 14 | 6 | 43.8 | 68.8 | 4.8 | 5.5 |
| 1007 | Proximal PER < REC < Distal PER | positive | 26 | 11 | 10 | 0 | 8 | 5 | 6.3 | 3.1 | 2.6 | 4.0 |
| 1008 | REC only (distal pudendal cut) | positive | 40 | 16 | 19 | 10 | 33 | 8 | 46.9 | 0.0 | 2.0 | 10.0 |
| 1010 | REC only (breathing artifact on P<sub>u</sub>) | positive | 40 | 19 | 39 | 32 | 33 | 13 | 75.0 | 81.3 | 3.6 | 7.2 |
| 1012 | Proximal PER / REC (no Distal PER) | positive | 31 | 16 | 0 | 0 | 16 | 19 | 50.0 | 56.3 | 20.3 | 18.9 |
| 1016 | REC < Distal PER < Proximal PER | positive | 35 | 30 | 16 | 19 | 26 | 26 | 53.1 | 50.0 | 22.8 | 2.8 |
| 1017 | Distal PER < REC < Proximal PER | positive | 24 | 21 | 12 | 12 | 15 | 8 | 40.6 | 46.9 | 2.0 | 3.2 |
| 1019 | REC < Distal PER < Proximal PER | positive | 24 | 18 | 17 | 19 | 21 | 15 | 56.3 | 50.0 | 6.4 | 4.6 |
| 1020 | Proximal PER < REC < Distal PER | explant | 36 | 36 | 30 | 26 | 24 | 22 | 71.9 | 40.6 | 6.1 | 2.8 |
| 1021 | REC < Proximal PER < Distal PER | positive | 26 | 5 | 15 | 10 | 21 | 6 | 37.5 | 37.5 | 3.2 | 2.6 |
| 1022 | Proximal PER / Distal PER < REC | positive | 35 | 6 | 37 | 6 | 27 | 4 | 37.5 | 12.5 | 14.5 | 15.9 |
| 1024 | REC < Proximal PER ~ Distal PER | positive | 23 | 11 | 6 | 0 | 16 | 7 | 18.8 | 3.1 | 8.5 | 24.3 |
| Mean ± SD |  |  | 29.8 ± 6.6 | 16.2 ± 9.2 | 16.8 ± 12.3 | 11.1 ± 10.3 | 20.2 ± 8.3 | 10.8 ± 7.5 | 44.0 ± 18.8 | 38.0 ± 26.0 | 7.8 ± 7.0 | 8.4 ± 7.0 |
| Paired t-test |  |  | p = 0.0001 |  | p = 0.0375 |  | p = 0.0026 |  | p = 0.283 |  | p = 0.797 |  |

Four participants exhibited a dominant proximal perineal nerve response (1007,1012, 1020, and 1021) while two participants had a dominant distal response (1016, 1017). Four participants had mixed responses to different electrodes (1001, 1019, 1022, 1024). Participant 1002 had all urethra sensors follow the same onset pattern (Table S2).

Out of 13 participants, 11 had positive implant outcomes (Table 2). Overall, participants had statistically significant improvement in pain (fGUPI) and bladder symptoms (Incontinence – M-ISI; Bladder – AUASI) after the PNS implant. There was no clear trend for bowel symptoms (CRAD-8) or sexual function (FSFI), which were not part of the initial symptoms for which participants were receiving the neurostimulator (Table 1). The order of recruitment for the perineal or rectal nerve did not have a relationship to symptom changes based on the overlap in 95% confidence intervals for each recruitment group (Table S3).

## Discussion

This was the first study to collect urethral pressure responses during pudendal neurostimulator implant surgeries. The intraoperative data collection allowed us to observe real-time stimulation-driven urethral responses, in addition to EAS EMG, and provided subject-specific pudendal nerve recruitment orders. The various pudendal nerve recruitment orders across participants (Table 2, Table S2) suggests a need to further investigate urethral activation patterns during pudendal nerve stimulation, which may help guide subject-specific stimulator programming and electrode placement.

We used a multi-sensor catheter to measure semi-continuous pressure in the lower urinary tract while participants were sedated in a prone position. Our baseline bladder pressure measurements (Figure S3) were consistent with prior studies that had similar participant age ranges (18-24 cmH_2_O)^21,22^. However, our maximum baseline urethral pressures differed from prior studies, which may have been related to the participant position. Previous studies have reported a maximum urethral pressure ranges of 68-84 cmH_2_O in a semi-seated position^21^ and 67-74 cmH_2_O in a semi-lithotomy position^22^, as compared to our average maximum measurement of 57 cmH_2_O (range 41-86 cmH_2_O, without an outlier). The prone position used in our study, which is least influenced by gravity, results in a lower pressure placed on the pelvic floor. This aligns with the established understanding that position can affect urethral pressure profiles^23^ and is the likely factor in the lower resting MUCP observed in our study (32 cmH_2_O) as compared to a prior study with the Manoscan catheter (59 cmH_2_O)^14^. Age, bladder filling state, and bladder underlying condition also all contribute to differences in urethral pressure profile^21-24^ and may have been a factor here.

Of our thirteen participants, seven were found to have the rectal nerve recruited first (Table 2 and Table S2). EAS EMG is a primary indicator of lead position and our observation that many participants had the rectal nerve recruited first is not surprising. Still, four participants had the perineal nerve recruited before the rectal nerve across rectal-responsive electrodes and the other two participants had a specific electrode with a perineal response first (Table S2). These findings imply that perineal nerve responses could serve as a useful indicator for pudendal implant electrode placement. In support of this, participant 1022, who lacked EAS responses at E3 within our tested amplitudes (Table S1), had perineal urethral responses (Table S2). The selective activation of the perineal nerve in this participant might be attributable to the E3 electrode in proximity to the pudendal nerve after the branching of the rectal nerve. Further investigation focusing on the variability of pudendal nerve branching anatomy and the relative positions of implanted electrodes to the nerves are necessary for understanding PNS activation patterns.

We observed that increases in stimulation amplitude above EASt did not elicit proportional increases in urethral responses (Figure 2). In seven participants, twice EASt stimulation amplitudes generated 66-100% of maximum urethral responses. Intriguingly, urethral responses decreased in a few participants when EASt amplitudes increased beyond threefold of EASt. This observation suggests that amplitudes above twice EASt may not trigger proportional urethral responses. A recent study with SNM patients presented similar findings: an increase in stimulation amplitude from 2x to 5x motor threshold amplitude only induced a 5% increase in sacral evoked responses^25^. These observations may have implications for device programming guidelines, as high stimulation amplitude may result in discomfort and unintentional activation of non-target nerves. A more extensive study investigating how participants with pudendal nerve implants utilize their devices at home may provide valuable insights into any relationships between stimulation settings and patients’ sensations.

There were a few limitations to consider in our PNS study. First, our study participant group was not racially or ethnically diverse. This was due to the pool of patients referred to the treatment plan from our collaborating clinic. Second, our data collection was confined to the duration of the pudendal implant surgeries. To ensure minimal interruption to the surgical procedures, we did not evaluate all potential electrode combinations or stimulation parameters.

## Conclusion

This study showed that female participants with pelvic pain and bladder symptoms had positive outcomes from the pudendal neuromodulation. There was no distinct correlation between the initial activation of either the perineal or rectal nerves and the change in bladder or pain symptoms. By monitoring stimulation-driven urethral responses intraoperatively, which are neglected in the current clinical approach, we were able to explore the pudendal nerve recruitment order. The recruitment order could assist in tailoring stimulation programs and parameters to individual patient needs, thereby enhancing treatment success. Altogether, this study broadened our understanding of pudendal neuromodulation by investigating intraoperative urethral responses and nerve activation patterns.

## Data Availability

The data set from the study will be found on the SPARC Science data portal at DOI: 10.26275.pc8r-r3iu after completing the SPARC curation process.

https://doi.org/10.26275.pc8r-r3iu

## Acknowledgments

Our deepest gratitude goes out to all the participants of our study. We thank Mackenzie Moore and Lauren Madden for their help in one of the data collection sessions. We appreciate the efforts of clinical study coordinators Mackenzie Moore and Vanessa Pruitt in recruiting patients for our study. We are also thankful to the Urology clinical team members, Neurology intraoperative monitoring staff including Kelly Ridenour and Stephanie Jones, and Medtronic representatives Jeff Drummond and Meghan Roth for their invaluable assistance in the operation room. We are especially grateful to Dr. Jim Hokanson for a discussion about stimulation-driven pressure tone increase.

## References

1. Peters KM, Killinger KA, Boguslawski BM, Boura JA. Chronic pudendal neuromodulation: expanding available treatment options for refractory urologic symptoms. Neurourol Urodyn. Sep 2010;29(7):1267–71. doi:10.1002/nau.20823

2. Peters KM, Feber KM, Bennett RC. A prospective, single-blind, randomized crossover trial of sacral vs pudendal nerve stimulation for interstitial cystitis. BJU Int. Oct 2007;100(4):835–9. doi:10.1111/j.1464-410X.2007.07082.x

3. Spinelli M, Malaguti S, Giardiello G, Lazzeri M, Tarantola J, Van Den Hombergh U. A new minimally invasive procedure for pudendal nerve stimulation to treat neurogenic bladder: description of the method and preliminary data. Neurourol Urodyn. 2005;24(4):305–9. doi:10.1002/nau.20118

4. Peters KM, Killinger KA, Jaeger C, Chen C. Pilot Study Exploring Chronic Pudendal Neuromodulation as a Treatment Option for Pain Associated with Pudendal Neuralgia. Low Urin Tract Symptoms. Sep 2015;7(3):138–42. doi:10.1111/luts.12066

5. Killinger KA, Kangas JR, Wolfert C, Boura JA, Peters KM. Secondary changes in bowel function after successful treatment of voiding symptoms with neuromodulation. Neurourol Urodyn. Jan 2011;30(1):133–7. doi:10.1002/nau.20975

6. Huang J, Fan Y, Zhao K, et al. Comparative Efficacy of Neuromodulation Technologies for Overactive Bladder in Adults: A Network Meta-Analysis of Randomized Controlled Trials. Neuromodulation. Dec 2023;26(8):1535–1548. doi:10.1016/j.neurom.2022.06.004

7. Bartley J, Gilleran J, Peters K. Neuromodulation for overactive bladder. Nat Rev Urol. Sep 2013;10(9):513–21. doi:10.1038/nrurol.2013.143

8. Maldonado PA, Chin K, Garcia AA, Corton MM. Anatomic variations of pudendal nerve within pelvis and pudendal canal: clinical applications. Am J Obstet Gynecol. Nov 2015;213(5):727.e1-6. doi:10.1016/j.ajog.2015.06.009

9. Montoya TI, Calver L, Carrick KS, Prats J, Corton MM. Anatomic relationships of the pudendal nerve branches. Am J Obstet Gynecol. Nov 2011;205(5):504 e1–5. doi:10.1016/j.ajog.2011.07.014

10. Gustafson KJ, Zelkovic PF, Feng AH, Draper CE, Bodner DR, Grill WM. Fascicular anatomy and surgical access of the human pudendal nerve. World J Urol. Dec 2005;23(6):411–8. doi:10.1007/s00345-005-0032-4

11. Barber MD, Bremer RE, Thor KB, Dolber PC, Kuehl TJ, Coates KW. Innervation of the female levator ani muscles. Am J Obstet Gynecol. Jul 2002;187(1):64–71. doi:10.1067/mob.2002.124844

12. Mahakkanukrauh P, Surin P, Vaidhayakarn P. Anatomical study of the pudendal nerve adjacent to the sacrospinous ligament. Clin Anat. Apr 2005;18(3):200–5. doi:10.1002/ca.20084

13. Peters KM. Pudendal neuromodulation for sexual dysfunction. J Sex Med. Apr 2013;10(4):908–11. doi:10.1111/jsm.12138

14. Kirby AC, Tan-Kim J, Nager CW. Dynamic maximum urethral closure pressures measured by high-resolution manometry increase markedly after sling surgery. Int Urogynecol J. Jun 2015;26(6):905–9. doi:10.1007/s00192-014-2622-4

15. Knowles CH, de Wachter S, Engelberg S, et al. The science behind programming algorithms for sacral neuromodulation. Colorectal Dis. Mar 2021;23(3):592–602. doi:10.1111/codi.15390

16. Clemens JQ, Calhoun EA, Litwin MS, et al. Validation of a modified National Institutes of Health chronic prostatitis symptom index to assess genitourinary pain in both men and women. Urology. Nov 2009;74(5):983-7.e3. doi:10.1016/j.urology.2009.06.078

17. Barry MJ, Fowler FJ, Jr., O’Leary MP, et al. The American Urological Association symptom index for benign prostatic hyperplasia. The Measurement Committee of the American Urological Association. J Urol. Nov 1992;148(5):1549–57. doi:10.1016/s0022-5347(17)36966-5

18. Suskind AM, Dunn RL, Morgan DM, DeLancey JO, McGuire EJ, Wei JT. The Michigan Incontinence Symptom Index (M-ISI): a clinical measure for type, severity, and bother related to urinary incontinence. Neurourol Urodyn. Sep 2014;33(7):1128–34. doi:10.1002/nau.22468

19. Barber MD, Walters MD, Bump RC. Short forms of two condition-specific quality-of-life questionnaires for women with pelvic floor disorders (PFDI-20 and PFIQ-7). Am J Obstet Gynecol. Jul 2005;193(1):103–13. doi:10.1016/j.ajog.2004.12.025

20. Rosen R, Brown C, Heiman J, et al. The Female Sexual Function Index (FSFI): a multidimensional self-report instrument for the assessment of female sexual function. J Sex Marital Ther. Apr-Jun 2000;26(2):191–208. doi:10.1080/009262300278597

21. Rud T. Urethral pressure profile in continent women from childhood to old age. Acta Obstet Gynecol Scand. 1980;59(4):331–5. doi:10.3109/00016348009154090

22. Henriksson L, Andersson KE, Ulmsten U. The urethral pressure profiles in continent and stress-incontinent women. Scand J Urol Nephrol. 1979;13(1):5–10. doi:10.3109/00365597909179993

23. Henriksson L, Ulmsten U, Andersson KE. The effect of changes of posture on the urethral closure pressure in healthy women. Scand J Urol Nephrol. 1977;11(3):201–6. doi:10.3109/00365597709179952

24. Dahms SE, Lampel DS, Kloeppel S, et al. Low urethral pressure profile--clinical implications. Scand J Urol Nephrol Suppl. 2001;35(207):100–5.

25. Goudelocke C, Jungbauer Nikolas LM, Bittner KC, Offutt SJ, Miller AE, Slopsema JP. Sensing in Sacral Neuromodulation: A Feasibility Study in Subjects With Urinary Incontinence and Retention. Neuromodulation. Feb 2024;27(2):392–398. doi:10.1016/j.neurom.2023.07.002

