## Supplemental for "Perineal and rectal nerve recruitment order varies during pudendal neurostimulator implant surgery"

### Supplemental Information

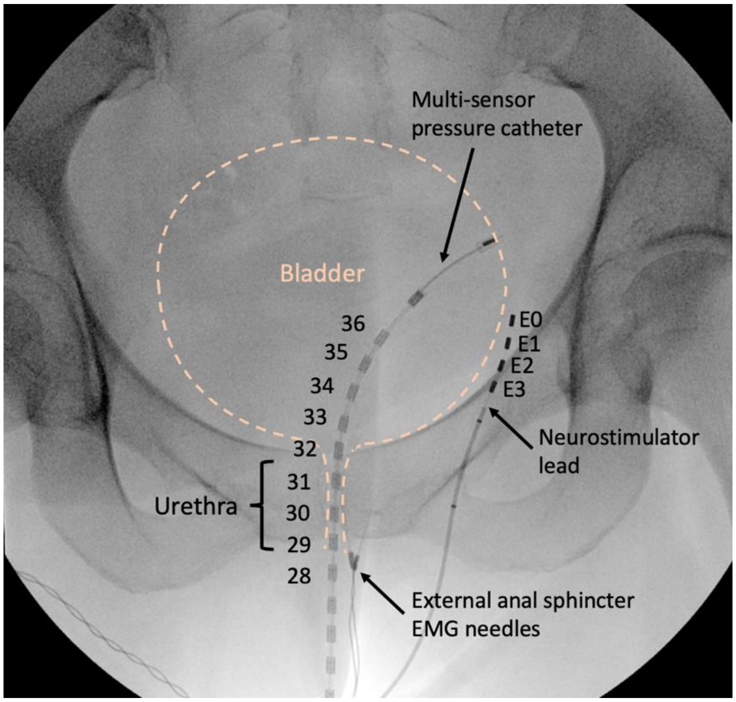


**Supplementary Figure S1.** Intraoperative fluoroscopy image showing PNS lead and Manoscan pressure catheter. The electrodes on the PNS lead are labelled E0-E3. The pressure catheter has thirty-six sensors, with the distal locations labelled here. Also indicated are the standard EAS EMG needles. The dashed line is an approximation of the bladder and urethra soft tissue for illustration purposes.

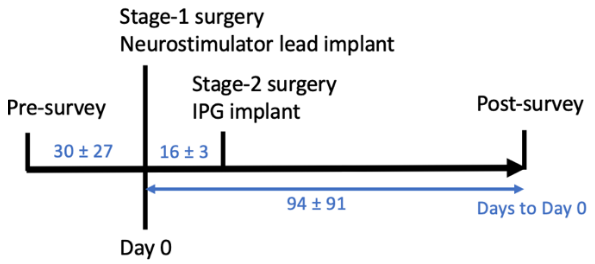


| | Pre-Survey | IPG Implant | Post-Survey |
|---|---|---|---|
| **1001** | -21 | +14 | +369 |
| **1002** | -46 | +21 | +110 |
| **1007** | -82 | +12 | +127 |
| **1008** | -29 | +16 | +34 |
| **1010** | -16 | +19 | +151 |
| **1012** | R | +16 | +51 |
| **1016** | -44 | +0 | +88 |
| **1017** | -20 | +0 | +36 |
| **1019** | -70 | +21 | +48 |
| **1020** | -4 | +14 | +42 |
| **1021** | -0 | +0 | +63 |
| **1022** | R | +14 | +57 |
| **1024** | -2 | +14 | +46 |
| **Mean ± SD** | 30 ± 27 | 16 ± 3 | 94 ± 91 |

**Supplementary Figure S2.** Timeline for participants in the study, in days. Two participants completed their surveys retrospectively (R), based on symptoms before PNS implant. Implant revision participants had a same day stage-1 and stage-2 combined surgery (+0). These two conditions were excluded from the summary statistics (grey).

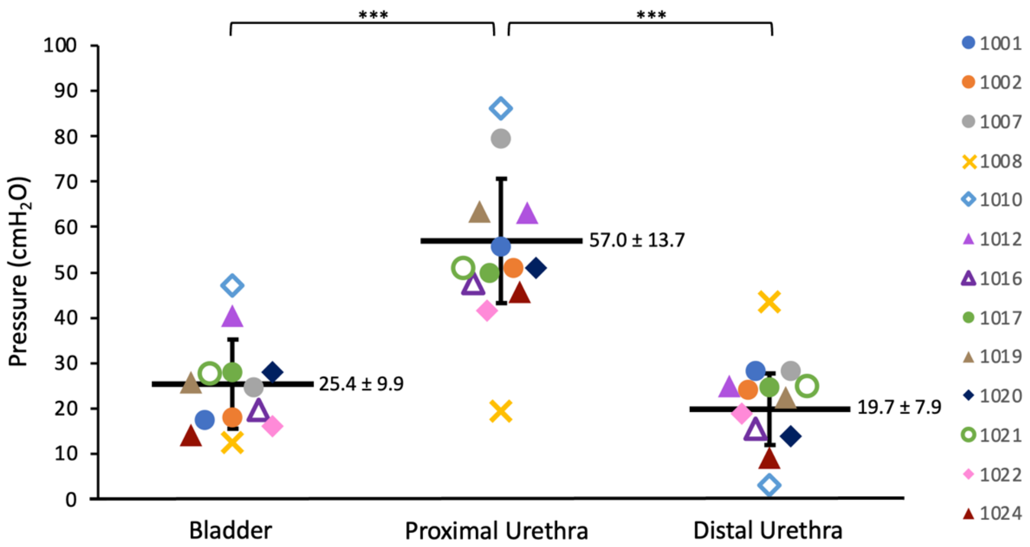


**Supplementary Figure S3.** Baseline lower urinary tract pressures for participants before initiation of stimulation trials. The Outlier, participant 1008 (yellow x), was excluded from summary statistics. This participant had a prior partial distal pudendal nerve resection due to a non-bladder-related medical condition, leading to different pressure trends from other participants. *** indicates $p < 0.0001$.

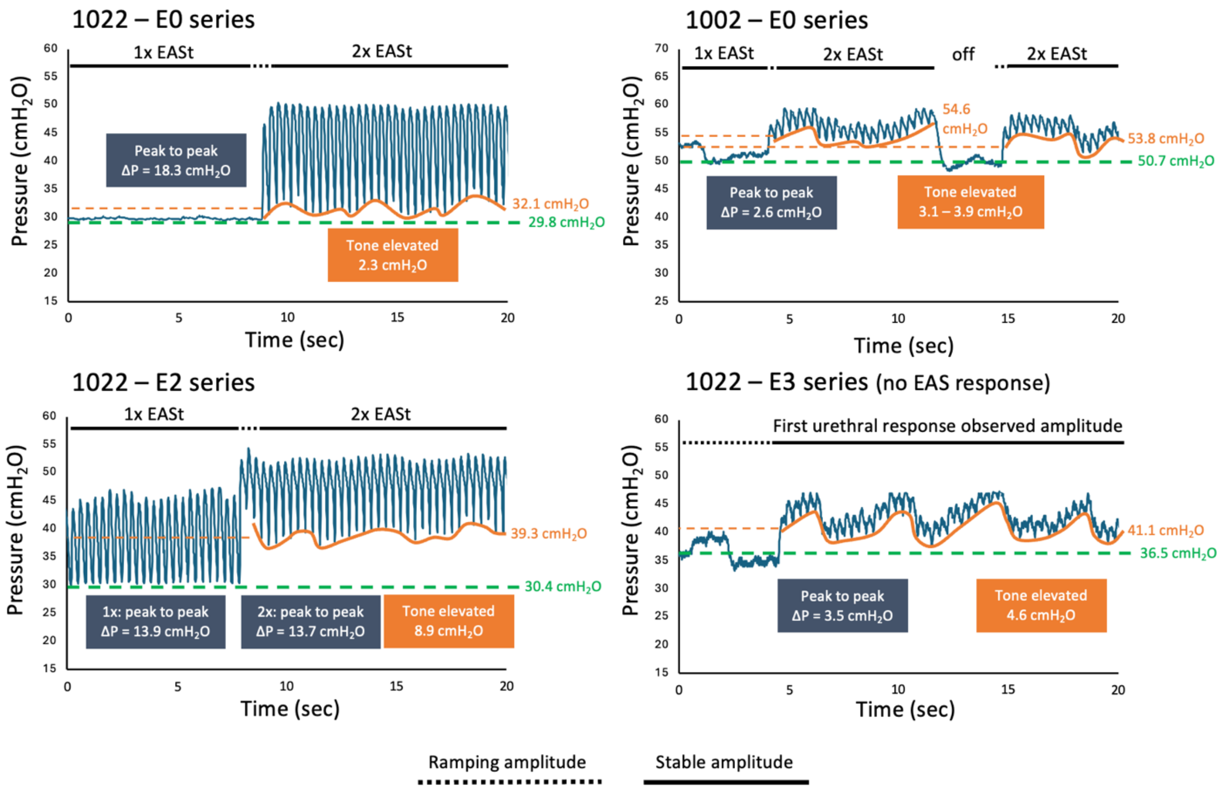


**Supplementary Figure S4.** Examples of stimulation-driven elevated pressure tone in the proximal urethra are shown for transitions in stimulation amplitude. Green dashed lines indicate the baseline pressure. Orange solid lines indicate the approximate pressure baseline during the elevated period. Orange dashed lines indicate the mean elevated pressure tone. Dark blue boxes represent the peak-to-peak ΔP of each amplitude, which are independent from the baseline tone increase.

**Supplementary Table S1.** External anal sphincter EMG motor threshold summary across participants.

| **Participant** | **E0** (mA) | **E1** (mA) | **E2** (mA) | **E3** (mA) |
|---|---|---|---|---|
| **1001** | 0.4 | 1.2 | 2.9 | x |
| **1002** | 1.5 | 0.5 | 0.4 | 2.1 |
| **1007** | 1.2 | 1.2 | 2.8 | x |
| **1008** | 1.0 | 1.1 | 1.1 | 1.4 |
| **1010** | 1.4 | 1.7 | 2.3 | x |
| **1012** | x | 4.1 | 3.1 | x |
| **1016** | 1.4 | 1.2 | 4.0 | 3.1 |
| **1017** | 1.1 | 2.0 | 0.8 | 3.0 |
| **1019** | 0.7 | 0.8 | 2.3 | x |
| **1020** | 1.1 | 1.0 | 1.1 | 2.7 |
| **1021** | 0.7 | 0.7 | 0.7 | 0.7 |
| **1022** | 0.7 | 2.4 | 1.6 | x |
| **1024** | 1.7 | 0.8 | 1.2 | 1.5 |
| Mean ± SD (N) | 1.1 ± 0.4 (12) | 1.4 ± 1.0 (13) | 1.9 ± 1.1 (13) | 2.1 ± 0.9 (7) |
| Median | 1.1 | 1.2 | 1.6 | 2.1 |

X - electrodes that did not elicit a response up to 5.5 mA and are not included in the summary statistics.

**Supplementary Table S2.** Urethral activation pattern for each participant.

| Electrode | **1001** | **1002** | **1007** | **1012** | **1016** | **1017** | **1019** | **1020** | **1021** | **1022** | **1024** |
|---|---|---|---|---|---|---|---|---|---|---|---|
| **E0** | D → All | All | P → All | XX | D → All | D → All | D | P → All | P | D → All | D |
| **E1** | All | All | P → All | P | D → All | D | All | P → All | P | All | P → All |
| **E2** | All | All | P | P | X | D | All | P → All | X | All | All |
| **E3** | XX | X | XX | XX | D | D | XX | P → All | X | P | P |

Proximal urethra (P), distal urethra (D), or among all 3-4 urethral sensors (ALL) indicates response to stimulation occurred. Arrow (→) indicates that as the amplitude was increased, the activation pattern expanded beyond the original location. The cells in grey indicate that a urethral response was captured before rectal nerve activation. Single X (X) indicates the rectal nerve was captured without perineal nerve responses. Double X (XX) indicates the electrode did not drive rectal or perineal nerve responses.

**Supplementary Table S3.** 95% confidence intervals for change in each survey score based on population standard deviation.

| Nerve recruitment | fGUPI | M-ISI | AUASI | CRAD-8 | FSFI |
|---|---|---|---|---|---|
| Rectal nerve first (N=7) | (-21.4, -8.6) | (-10.8, 2.2) | (-18.4, -5.3) | (-20.1, 8.5) | (-5.0, 6.1) |
| Perineal nerve first (N=4) | (-20.3, -3.2) | (-19.9, -2.6) | (-17.5, 0.0) | (-32.2, 5.6) | (-7.6, 7.9) |
| Mixed nerve activation (N=2) | (-24.6, -0.4) | (-11.7, 12.7) | (-13.8, 10.8) | (-18.9, 34.5) | (-10.3, 11.5) |
